# Genetic and Epigenetic Regulation of Inflammatory Genes Drives Calcific Aortic Valve Disease (CAVD)

**DOI:** 10.1101/2025.05.15.25327710

**Authors:** Shu-Ying Ding, Rena Rehemuding, Xiao-Lei Li, Xiao-Mei Li, Xian Wei, Zi-Xiang Yu, Fen Liu, Munawaer Keremu, Jun Cui, Xia Li, Dilare Adi, Yi-Tong Ma

## Abstract

**Introduction:** Calcific aortic valve disease (CAVD) is the most common valvular disorder in older adults, characterized by progressive fibrosis, calcification, and impaired blood flow. Despite its growing burden, no effective pharmacological treatments exist. Inflammation and epigenetic dysregulation are increasingly recognized as central to CAVD pathogenesis. This study aimed to identify key inflammatory genes and regulatory mechanisms contributing to disease development.

**Methods:** We performed a meta-analysis of aortic valve transcriptomic datasets, focusing on inflammation-related genes, to identify differentially expressed genes (DEGs) in CAVD. Blood-derived expression quantitative trait loci (eQTL) and DNA methylation QTL (mQTL) data were integrated with CAVD genome-wide association study (GWAS) results (FinnGen database) using a three-step summary data-based Mendelian randomization (SMR) framework. Additionally, we combined aortic eQTL data and inflammatory mediator GWAS to assess tissue-specific regulatory interactions through SMR and co-localization analyses.

**Results:** Fifty-seven inflammation-related DEGs were identified, enriched in immune cells from patients with CAVD. Multi-omics integration prioritized nine causal genes in blood, including PPARG, TLR2, LTA, TNF, C4A, IL6, RELA, C3, and TGFβ1. Among these, TNF and TGFβ1 were strongly associated with increased CAVD risk. In aortic tissue, SMR and co-localization analyses revealed TLR2, C4A, and AGER as key genes linked to inflammatory pathways, highlighting a potential gene-immune axis in disease progression.

**Conclusions:** Our integrative multi-omics analysis reveals that calcific aortic valve disease (CAVD) is driven by genetically and epigenetically regulated inflammatory pathways. We identify TNF and TGFβ1 as epigenetically controlled causal drivers, while AGER, C4A, and TLR2 emerge as tissue-specific mediators of aortic inflammation. These findings establish a causal link between immune dysregulation and CAVD pathogenesis, highlighting promising targets for biomarker development and precision therapies aimed at interrupting disease progression.

## 1. Introduction

Calcific aortic valve disease (CAVD) is a progressive fibro-calcific disorder that affects the aortic valve, ultimately leading to stenosis and impaired cardiac function. It is the second most common cardiovascular disease after coronary artery disease and hypertension [1,2], with a prevalence rising from 20%-30 % in individuals over 65 years to nearly 50%-57% in those older than 85 years [3]. Despite its growing clinical burden, no effective pharmacological therapies exist to halt or slow disease progression, and valve replacement remains the only definitive treatment [4].

Once considered a passive degenerative consequence of aging, CAVD is now recognized as an active, cell-mediated process involving chronic inflammation, immune cell infiltration, and osteogenic differentiation of valve interstitial cells (VICs) [5]. Its pathological parallels with atherosclerosis (AS), including lipid accumulation, endothelial dysfunction, and persistent inflammation, suggest shared mechanisms [6]. Histological analyses have revealed inflammatory cell infiltration in up to 28% of calcified valves [7], and elevated levels of pro-inflammatory cytokines such as interleukin-1β (IL-1β) and tumor necrosis factor-alpha (TNF-α) have been implicated in promoting VIC activation and calcification [8,9]. Additionally, systemic inflammatory indices, such as neutrophil-to-lymphocyte and platelet-to-lymphocyte ratios, have been correlated with disease severity [10–12], suggesting that CAVD reflects both localized and systemic inflammatory dysregulation [13–15]. Although observational and experimental data underscore a strong association between inflammation and CAVD, causality remains uncertain. Emerging evidence also points to the role of epigenetic modifications, particularly DNA methylation, in regulating inflammatory gene expression and disease susceptibility [16].

Given these insights, we hypothesized that inflammation-related genes, modulated by genetic and epigenetic factors, contribute causally to CAVD pathogenesis. To investigate this, we performed an integrative multi-omics analysis combining transcriptomic meta-analysis, genome-wide association study (GWAS) data, and expression and methylation QTL analyses, including tissue-specific evaluations from the aortic valve. Our goal was to delineate key inflammatory pathways and regulatory mechanisms underlying CAVD and identify novel therapeutic targets.

## 2. Methods

### 2.1 Study design and data resources

This study aimed to identify inflammation-related genes implicated in calcific aortic valve disease (CAVD) through multi-omics integration. Candidate genes were selected from the GeneCards database based on an inflammation relevance score ≥10. Four publicly available transcriptomic datasets of aortic valve tissue (GSE12644, GSE51472, GSE83453, and GSE88803) were obtained from the Gene Expression Omnibus (GEO) [17], including samples from patients with CAVD and healthy controls. A meta-analysis using linear regression was performed to identify differentially expressed genes (DEGs), adjusting for multiple comparisons using the false discovery rate. To assess genetic regulation, DEGs were integrated with CAVD genome-wide association study (GWAS) summary statistics from the FinnGen consortium (12,194 cases, 28,072 controls). Blood-derived expression quantitative trait loci (eQTL) data from the eQTLGen consortium (n = 31,684) [18] and DNA methylation QTL (mQTL) data from the Brisbane Systems Genetics Study (n = 614) and Lothian Birth Cohort (n = 1,366) [19] were utilized to explore genetic and epigenetic influences. Tissue-specific eQTL data for the aorta (n = 860) were sourced from the GTEx project [20]. Additionally, GWAS data for circulating inflammatory mediators were retrieved from the GWAS Catalog [21]. Analyses were restricted to cis-eQTLs and cis-mQTLs (±1 Mb of target genes). Finally, the UK Biobank cohort was used to validate key findings. Detailed dataset descriptions and analytic workflows are provided in the Supplementary Materials.

### 2.2 Transcriptomic Meta-Analysis of Inflammation-Related Differentially Expressed Genes in CAVD

To identify inflammation-related genes involved in CAVD, we conducted a meta-analysis of differentially expressed genes (DEGs) using data from four independent aortic valve gene expression datasets. For each dataset, linear regression models were applied to compare gene expression between patients with CAVD and healthy controls (HCs), adjusting for available covariates such as age, sex, body mass index (BMI), and medication use. To account for variability in sample collection sites, tissue location was included as a covariate in each model. DEG analysis was performed separately for each dataset, after which results were combined using a fixed-effects meta-analysis implemented in R using the *metafor* package. This approach allowed us to improve statistical power while controlling for dataset-specific confounders. All clinical variables were standardized before analysis, and methodological rigor was applied to ensure reproducibility. As this was a secondary analysis of publicly available datasets, clinical trial registration was not required.

### 2.2 Mendelian Randomization and Co-localization Analyses to Identify Causal Inflammatory Drivers of CAVD

To identify causal inflammatory drivers of calcific aortic valve disease (CAVD), we conducted summary data-based Mendelian randomization (SMR) and co-localization analyses. For the blood-based analysis, we applied a three-step SMR approach: (1) single nucleotide polymorphisms (SNPs) were used as instrumental variables with gene expression as exposure and CAVD as outcome; (2) SNPs were tested with DNA methylation as exposure and CAVD as outcome; and (3) SNPs were evaluated with DNA methylation as exposure and gene expression as outcome, considering only signals significant in the first two steps. Genes were deemed putatively causal if they fulfilled all of the following: significant in all three SMR tests (false discovery rate [FDR] <0.05), genome-wide suggestive significance (P <1×10□□ for eQTLs, mQTLs, and GWAS), and no evidence of heterogeneity (HEIDI P >0.05), excluding horizontal pleiotropy. For the aortic tissue-based analysis, we performed SMR to evaluate associations between cis-eQTLs from aortic tissues (exposure) and CAVD GWAS data (outcome), prioritizing genes meeting SMR significance (FDR <0.05), genome-wide suggestive P-values (P <1×10□□), and no heterogeneity (HEIDI P >0.05). To explore shared regulatory mechanisms between inflammatory mediators and gene expression, we conducted co-localization analyses using the coloc R package, considering a posterior probability (PPH4) >0.5 as evidence for a shared causal variant. All statistical methods were performed following standard protocols to ensure reproducibility, and inclusion criteria encompassed only SNPs with strong cis-acting regulatory effects. No clinical trial registration was required, as this was a secondary data analysis of publicly available genomic datasets.

### 2.3 Validation of Inflammation-Related DEGs and Pathways in an Independent Multi-Omics Cohort

To validate our findings, we selected significantly differentially expressed genes (DEGs) identified through the meta-analysis of four aortic valve gene expression datasets. These DEGs were previously shown to distinguish patients with CAVD from healthy controls (HCs) and were replicated using data from the UK Biobank (UKBB). In addition, we examined key gene expression pathways associated with inflammatory factors that were identified through our co-localization analysis. These pathways were also assessed in the FAH-SYS IBD multi-omics cohort to further confirm their relevance to inflammation and CAVD pathogenesis.

### 2.4 Cell Type Enrichment and Epigenetic Annotation of Inflammation-Associated Loci

To investigate the cellular context of inflammation-related gene regulation, we performed cell-type-specific enrichment analysis using the Cell Type-Specific Enrichment Analysis Database (CSEA-DB, https://bioinfo.uth.edu/CSEADB/). Among 126 general cell types across 61 tissues, we focused on 27 cell types within the circulatory system. Multiple testing correction was applied using the Benjamini–Hochberg (BH) method, with significance set at *FDR* < 0.05.

We further examined the regulatory characteristics of DNA methylation sites using eFORGE (http://eforge.cs.ucl.ac.uk/). This tool was used to assess the enrichment of chromatin features like both active and inactive states, and histone modifications, specifically H3K4me1 and H3K4me3 marks. In addition, genomic regions surrounding the DNA methylation sites were annotated using the Ensembl Genome Browser (http://grch37.ensembl.org/).

### 2.5 Statistics

All analyses applied appropriate statistical methods with multiple testing corrections to ensure robust and reliable results. Differential expression in transcriptomic meta-analysis was assessed using effect size-based models, with significance defined as *FDR* < 0.05 (Benjamini–Hochberg correction). Cell type enrichment via CSEA was similarly corrected for multiple comparisons (*FDR* < 0.05). Summary-based Mendelian randomization (SMR) analyses integrated GWAS, eQTL, and mQTL data, with significance determined by *FDR* < 0.05, and validated by non-significant HEIDI (*P* > 0.05) and Cochran’s Q tests (*P* > 0.05) to exclude linkage and heterogeneity. eFORGE was used to test enrichment of chromatin states and histone marks at methylation sites, using empirical p-values and background-matched controls. Co-localization of eQTL and inflammatory QTL signals was evaluated by posterior probability (PPH4), with values > 0.5 considered suggestive and > 0.9 considered strong evidence of shared genetic architecture.

## Results

### 1. Meta-analysis Identifies Inflammation-Related DEGs in CAVD and Highlights Myeloid Cell Involvement

To investigate the contribution of inflammation to calcific aortic valve disease (CAVD), we performed a meta-analysis of four publicly available aortic valve transcriptome datasets comparing patients with CAVD (n = 30) and healthy controls (n = 37). From a curated set of 154 inflammation-related genes (GeneCards relevance score ≥10) as shown in Figure 1A, functional classification revealed enrichment for cytokines, chemokines, receptors, signaling molecules, and transcription factors, with top-ranked genes including NLRP3, IL6, TNF, CRP, IL10, TLR4 and IL1B, as shown in Figure 1B. Meta-analysis identified 57 differentially expressed genes (DEGs) (FDR < 0.05), including key mediators such as TNF, SPP1, PTGS2, HMGB1, and TGFB1, previously implicated in inflammatory signaling and tissue remodeling central to CAVD pathogenesis [22–26]. Cell-specific enrichment analysis (CSEA) demonstrated that these DEGs were predominantly enriched in myeloid populations, particularly macrophages and dendritic cells (FDR = 0.0024), as shown in Figure 1C. Highlighting the likely role of these immune cells in driving local inflammatory responses and promoting osteogenic reprogramming of valve interstitial cells in CAVD.

**Fig. 1.**
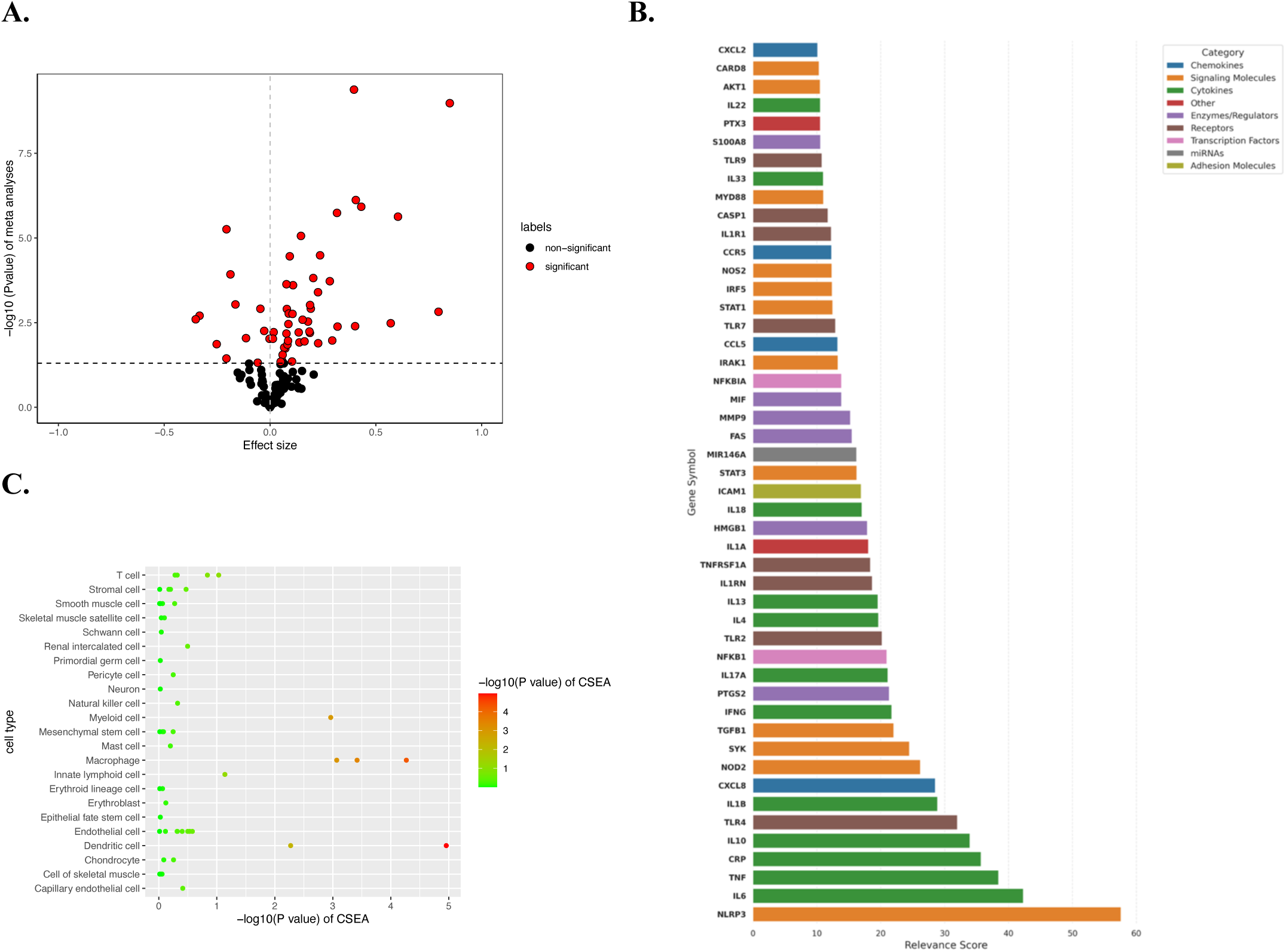
Inflammatory Gene Dysregulation and Immune Cell Enrichment Underpin CAVD Pathogenesis. (A) Volcano plot showing differential expression of 154 inflammation-related genes in CAVD, with 57 significantly altered (red dots; FDR < 0.05); the x-axis represents meta-effect sizes and the y-axis shows –log10-transformed meta-P values, with the dashed line marking the FDR threshold. (B) Horizontal bar plot categorizing inflammation-related genes by biological function, with relevance scores highlighting top mediators such as NLRP3, IL6, TNF, and CRP across cytokines, chemokines, receptors, and other groups. (C) Dot plot illustrating significant enrichment of inflammation-associated genes in specific valve- and blood-derived cell types, based on –log10(meta-P) values, emphasizing cellular contributors to CAVD pathology.

### 2. The integration of GWAS and blood eQTLs/mQTLs data reveals genes related to inflammation

To investigate the upstream regulation of inflammatory genes associated with CAVD, we performed a three-step Summary-based Mendelian Randomization (SMR) analysis integrating CAVD GWAS data with blood-derived cis-eQTL and cis-mQTL datasets. Given the role of DNA methylation in modulating gene expression, particularly in promoter and enhancer regions [19], we first utilized cis-eQTL data from the eQTLGen consortium (n = 31,684), identifying 11 inflammation-related genes significantly associated with CAVD (FDR < 0.05, HEIDI P > 0.05, Cochran’s Q P > 0.05; Supplementary Table S5). Extending this to epigenetic regulation, we combined GWAS data with cis-mQTL summary statistics from the Brisbane Systems Genetics Study and the Lothian Birth Cohort (n = 1,980), identifying 1,461 methylation probes near 127 candidate genes (±1 Mb; Supplementary Table S6). Integrated analysis of both cis-eQTL and cis-mQTL datasets subsequently prioritized nine genes with significant associations across all three SMR steps, as shown in Figure 2. (FDR < 0.05, HEIDI P > 0.05, Cochran’s Q P > 0.05; Supplementary Table S7), suggesting they are regulated through both genetic and epigenetic mechanisms. Collectively, these results implicate a model in which genetic variants and DNA methylation jointly modulate inflammation-related gene expression, contributing to CAVD susceptibility.

**Fig. 2.**
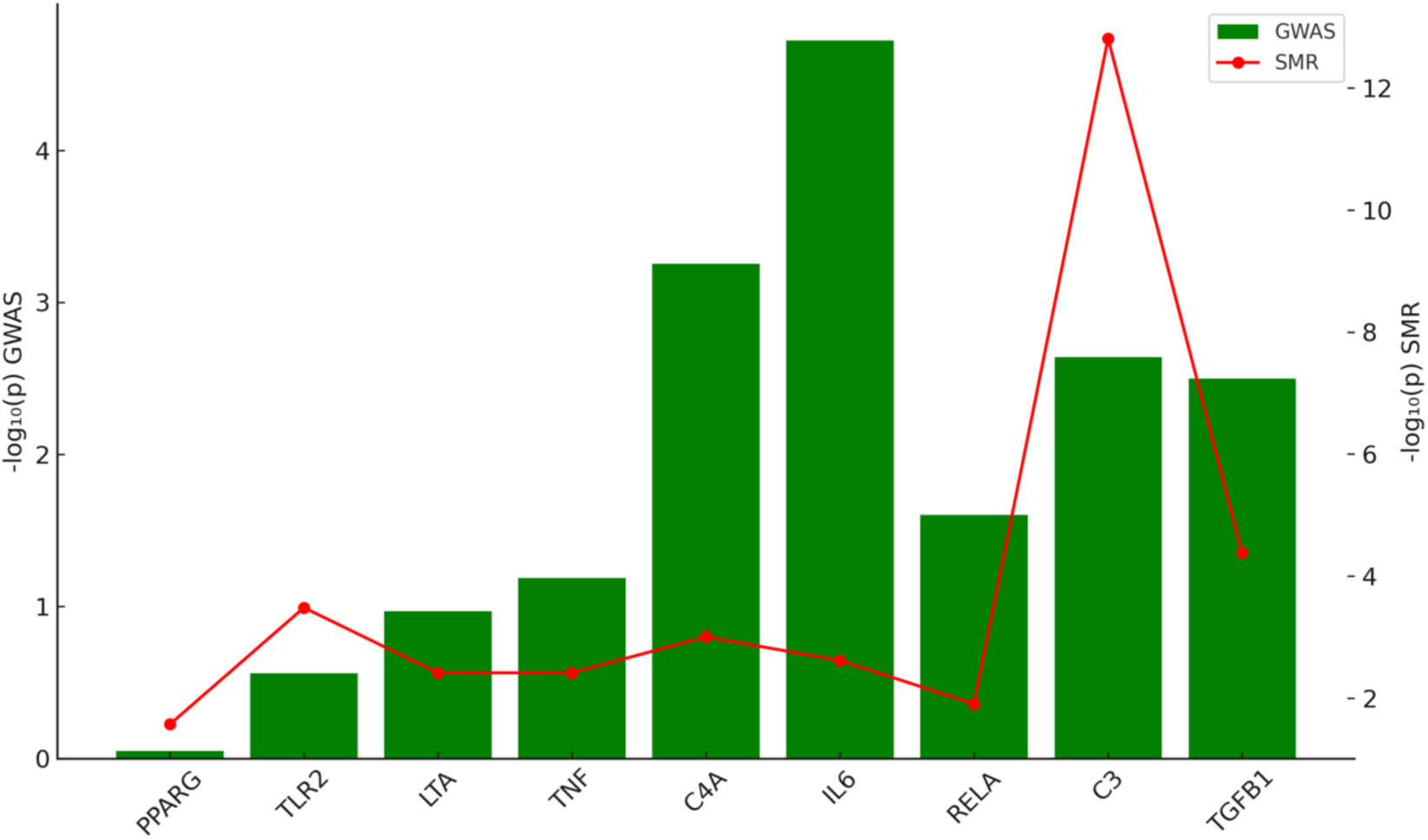
Integrated GWAS-SMR Analysis Identifies Causal Inflammatory Genes in CAVD. Comparison of genome-wide association study (GWAS) signals (green bars) and summary Mendelian randomization (SMR) results (red line) for key inflammation-related genes. The y-axis shows –log□□(p-value) on a logarithmic scale. This integrative analysis highlights genes with both strong genetic associations and regulatory effects, supporting their causal role in calcific aortic valve disease (CAVD) pathogenesis.

### 3. Blood-Based Methylation Regulation of Inflammatory Genes Highlights the Potential Role of TGFB1 and TNF in CAVD

Among the nine inflammation-related genes prioritized by integrative SMR analysis, TGFβ1 and TNF emerged as key candidates for further investigation based on the density and relevance of associated methylation probes and regulatory SNPs. Although C3 initially appeared promising, only a single SNP was identified, limiting causal inference. In contrast, TNF demonstrated a rich clustering of SNPs around enhancer and promoter regions, while TGFβ1 exhibited focused promoter-specific probes, suggesting biologically meaningful epigenetic regulation. Genes such as LTA and C4A showed broader SNP distributions, complicating functional interpretation. Prioritizing TGFβ1 and TNF, we found that promoter methylation at cg07663732 was positively associated with reduced TGFβ1 expression (βSMR = 0.102), implicating methylation-mediated transcriptional control in vascular remodeling and CAVD pathogenesis, as shown in Figures 3(A–B). Similarly, reduced methylation at cg01584389 near the TNF enhancer correlated with increased TNF expression (βSMR = –0.133) and heightened CAVD risk, as shown in Figures 3 (C–D), supporting a model whereby enhancer demethylation drives pro-inflammatory signaling in valve tissues. Together, these findings underscore the epigenetic regulation of TGFβ1 and TNF in blood and highlight their potential as molecular targets for early diagnostic or therapeutic strategies in CAVD.

**Fig. 3.**
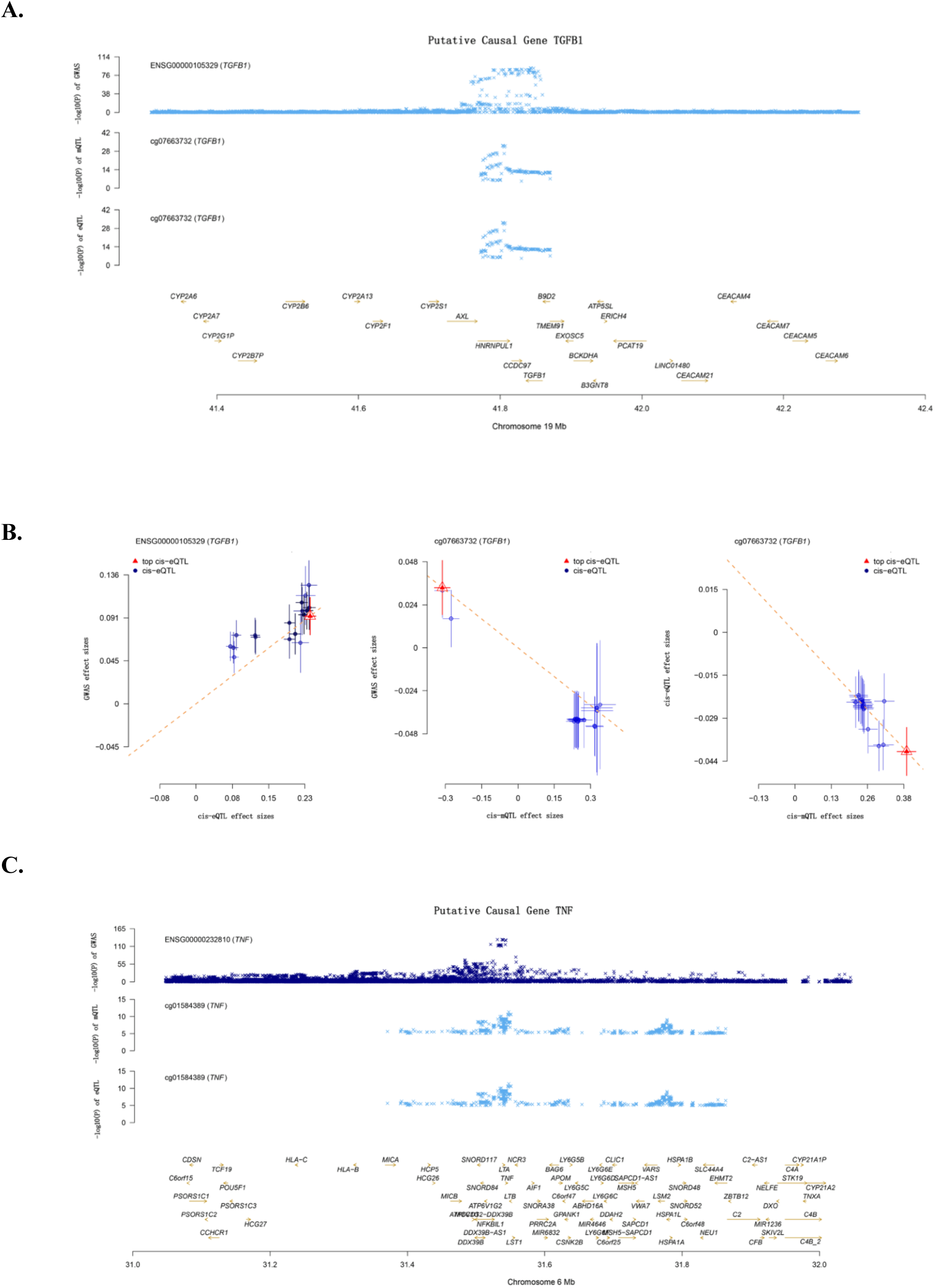
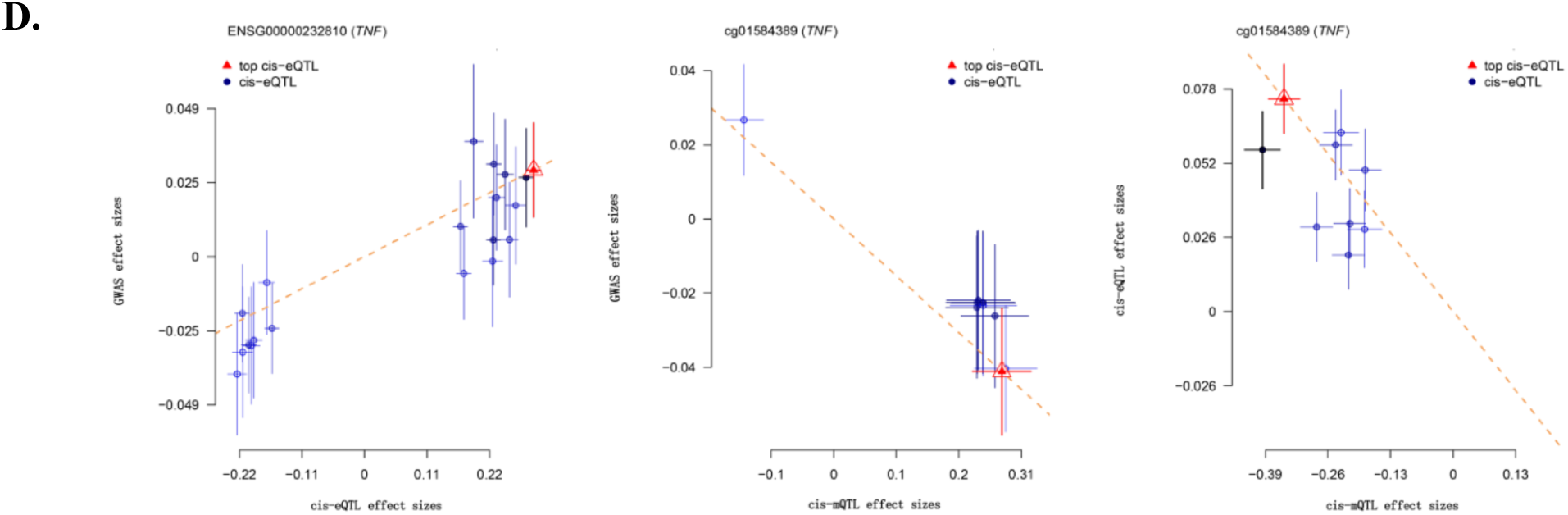
Epigenetic Regulation of TGFβ1 and TNF Drives Inflammatory Remodeling in CAVD. Panels A and C show the genetic association between cis-eQTLs near the TGFB1 and TNF loci and CAVD risk, highlighting key SNPs with P < 1 × 10□. Panels B and D illustrate the three-step SMR analysis, demonstrating significant causal pathways linking DNA methylation to gene expression and, ultimately, to CAVD susceptibility (FDR < 0.05; HEIDI P > 0.05). From left to right, plots depict: (1) gene expression versus CAVD GWAS association, (2) DNA methylation versus CAVD GWAS association, and (3) DNA methylation versus gene expression association. Together, these findings suggest that methylation-driven regulation of TGFB1 and TNF may promote inflammatory remodeling in valve tissues, contributing to CAVD pathogenesis.

### 4. Aortic Tissue eQTL Meta-Analysis Identifies Tissue-Specific Inflammatory Drivers of CAVD

Given the influence of tissue-specific regulatory mechanisms on disease susceptibility, and recognizing that gene expression is shaped by the unique microenvironment and physiological roles of the aorta, we next focused on identifying aortic-specific drivers of CAVD. Meta-analysis of aortic cis-eQTL data from the GTEx project (n = 860) revealed 43,975 significant SNP-gene associations involving 392 inflammation-related genes (FDR < 0.05; Supplementary Table S9). Summary data-based Mendelian randomization (SMR) analysis prioritized four genes namely TLR2, C4A, AGER, and CBS showing significant associations to CAVD (FDR < 0.05), each passing sensitivity tests for pleiotropy (HEIDI P > 0.05) and heterogeneity (Cochran’s Q P > 0.05), as shown in Figure 4 and detailed in Supplementary Table S10. To further investigate tissue-specific regulation, co-localization analysis was performed using matched QTL datasets for inflammatory mediators, revealing shared regulatory variants, particularly for AGER, C4A, and TLR2. These findings suggest that genetic variants at these loci may simultaneously influence aortic gene expression and systemic inflammation, offering mechanistic insights into the inflammatory underpinnings of CAVD pathogenesis (Supplementary Table S11) [27].

**Fig. 4.**
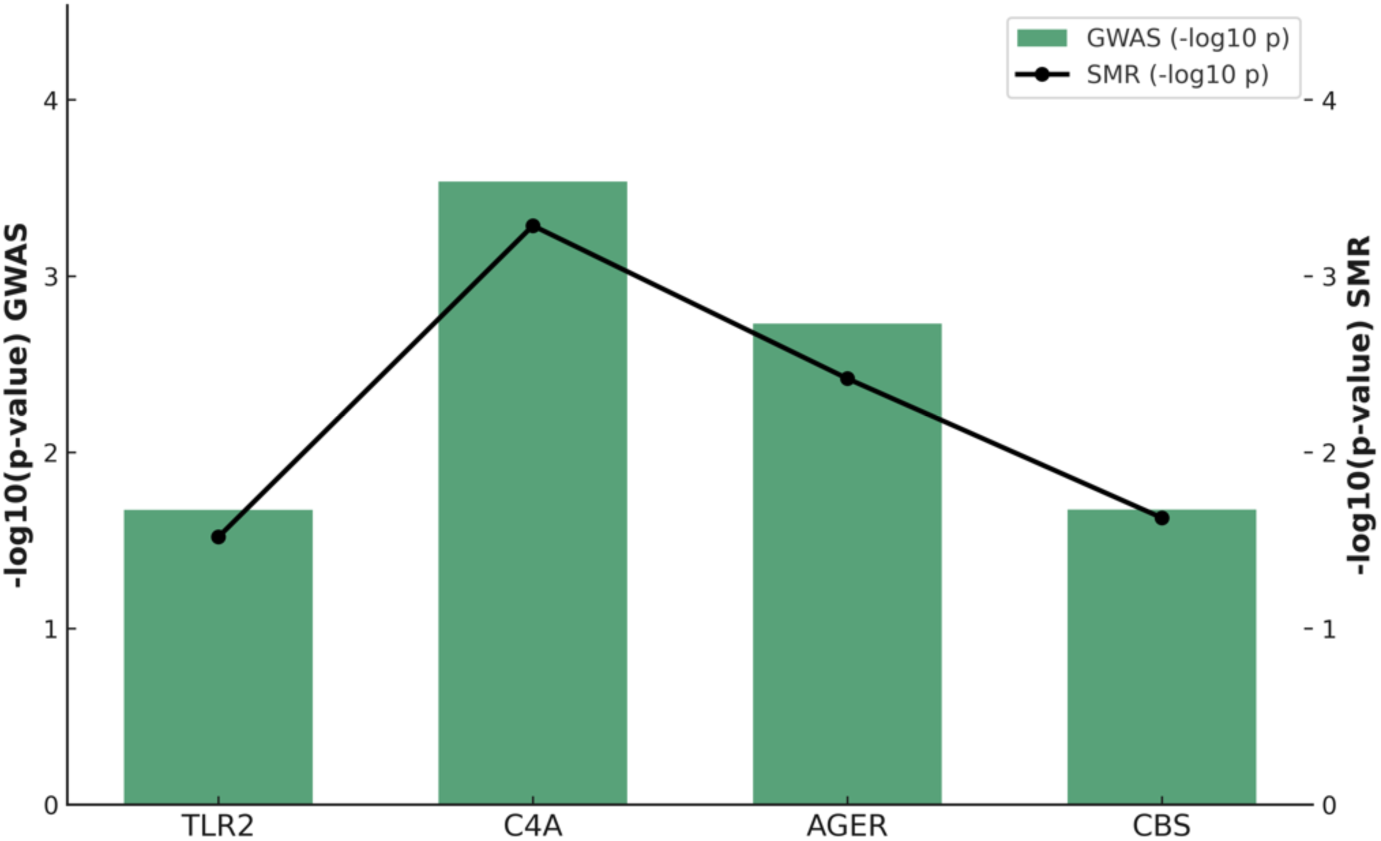
Aortic-Specific eQTL and GWAS Integration Reveals Key Inflammatory Gene Drivers of CAVD. Multi-omics integration of aortic cis-eQTL data with CAVD GWAS results identifies key candidate genes. Bar plots show –log□□(p) values from GWAS for lead SNPs near candidate loci, while the black line indicates –log□□(p) values from SMR analysis. TLR2, C4A, AGER, and CBS demonstrated significant SMR associations (FDR < 0.05) and passed HEIDI sensitivity testing (P > 0.05), suggesting a causal link between gene regulation and CAVD development.

### 5. Genetic Interactions Between Inflammatory Mediators and AGER, C4A, and TLR2 Reveal Mechanisms of Aortic Inflammation in CAVD

Using combined summary-based Mendelian randomization (SMR) and co-localization analyses, we identified AGER, C4A, and TLR2 as potential causal genes influencing inflammatory processes in aortic tissues affected by calcific aortic valve disease (CAVD). Increased AGER expression was positively associated with CAVD risk (β_SMR = 0.1858), with co-localization analyses revealing strong shared regulatory variants with inflammatory mediators such as SLAMF1 (PPHL = 0.9723) and CCL23 (PPHL = 0.9248), as shown in Figure 5A, which are implicated in immune activation and eosinophil differentiation [28]. Additional overlap was observed with IL-18 (PPHL = 0.6268) and IL-2RB (PPHL = 0.5072), mediators involved in amplifying T-cell responses and maintaining immune tolerance [29–30]. In contrast, higher C4A expression exerted a protective effect against CAVD (β_SMR = –0.0923), with co-localization identifying shared genetic regulation with nine inflammation-related mediators, including CD5, IL-15RA, CSF1, IL-17C, and others, shown in Figure 5B, thus highlighting C4A’s potential role in dampening inflammatory cascades [31]. Notably, TLR2 expression was positively associated with CAVD risk (β_SMR = 0.1685), and co-localized with IL-22RA1 (PPHL = 0.9476) and CXCL5 (PPHL = 0.9229), shown in Figure 5C, both known to drive pro-inflammatory signaling [32]. Together, these findings suggest that genetically regulated expression of AGER, C4A, and TLR2 modulates inflammatory pathways in the aorta, contributing to CAVD pathogenesis.

**Fig. 5.**
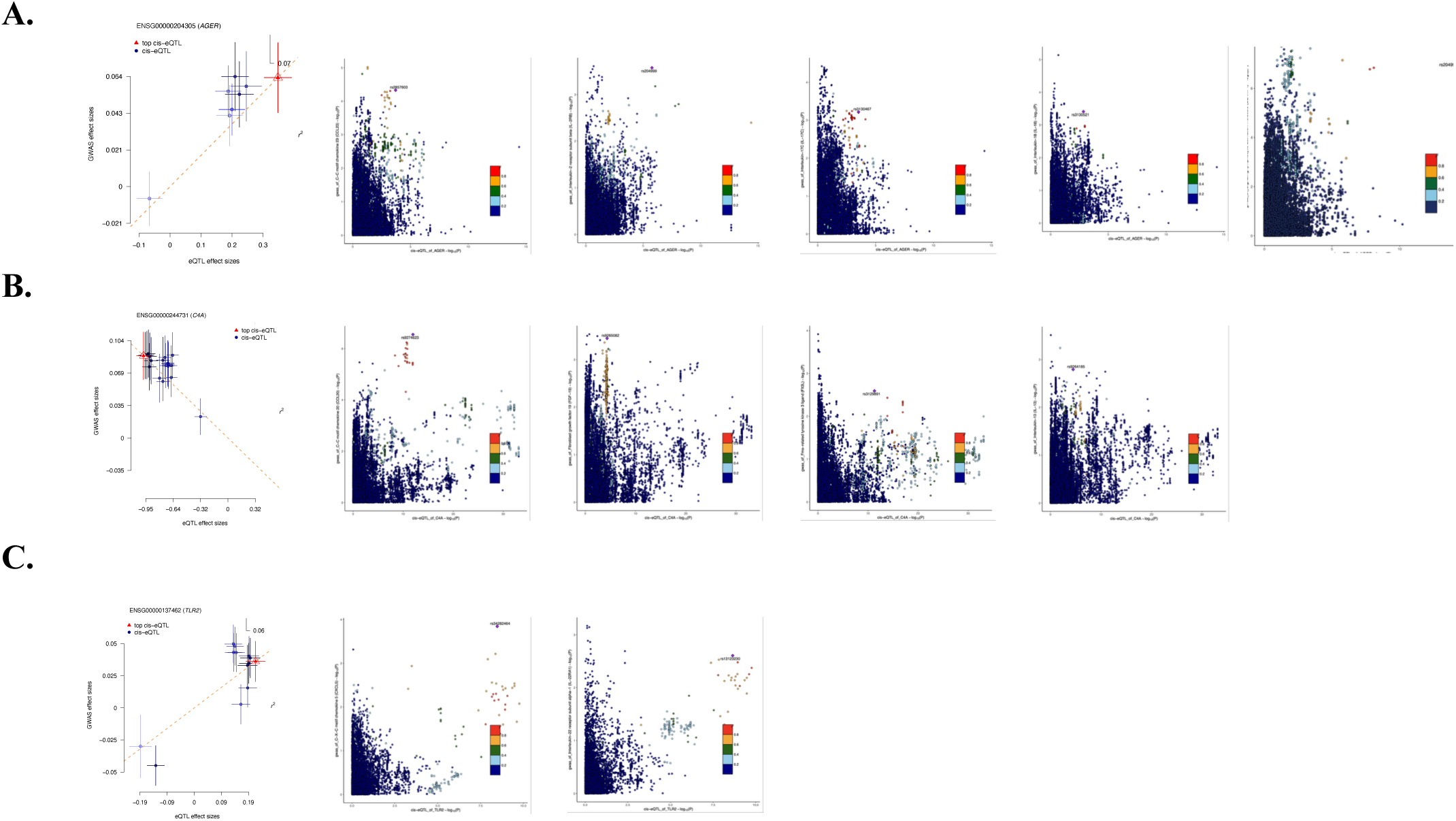
AGER, C4A, and TLR2 Mediate Aortic Inflammatory Pathways Linked to CAVD Risk. SMR and co-localization analyses identify AGER, C4A, and TLR2 as causal aortic genes associated with calcific aortic valve disease (CAVD) through inflammatory regulatory networks. The left panels depict SMR associations between gene expression and CAVD risk (all passing SMR FDR < 0.05; HEIDI P > 0.05), while the right panels illustrate co-localization between cis-eQTLs and mbQTLs for inflammatory mediators. r² values reflect linkage disequilibrium between variants and lead SNPs. Panels A–C correspond to AGER (A), C4A (B), and TLR2 (C), highlighting distinct inflammatory gene interactions contributing to CAVD pathogenesis.

### 6. Population analysis confirms Inflammatory genes as Genetic Risk Factors for CAVD

To validate the inflammation-related genes identified through Mendelian randomization (MR) analyses, we leveraged the large-scale UK Biobank (UKBB) cohort, encompassing a broad demographic spectrum (Supplementary Table S12). Key candidate genes were selected based on MR findings. Notably, the TNF locus (rs116454630) and the TGFβ1 locus (rs34503210) exhibited significant positive associations with CAVD risk, confirming their potential causal roles. Additional genes, including PPARG, TLR2, LTA, C4A, IL6, RELA, C3, AGER, and CBS, demonstrated consistent directional trends in association with CAVD, mirroring the results from MR analyses. These findings provide independent validation in a large population-based cohort and strengthen the evidence linking inflammation-related gene dysregulation to CAVD pathogenesis.

## Discussion

In this study, we leveraged a multi-omics framework to elucidate the inflammatory and regulatory mechanisms contributing to calcific aortic valve disease (CAVD). Our findings provide convergent evidence supporting the central role of inflammation in CAVD pathogenesis, while also revealing novel gene-mediator interactions with potential mechanistic and translational relevance. Through a meta-analysis of publicly available aortic valve transcriptomic datasets, we identified 57 inflammation-related differentially expressed genes (DEGs). Integrating these DEGs with GWAS, eQTL, and mQTL datasets, we identified nine candidate genes, namely, PPARG, TLR2, LTA, TNF, C4A, IL6, RELA, C3, and TGFβ1, as likely contributors to disease susceptibility. Notably, aortic-specific eQTL analysis further identified AGER, C4A, and TLR2 as tissue-specific contributors, implicating that both local vascular and systemic inflammatory pathways are involved in CAVD development. These findings support the existing evidence that chronic inflammation leads to valvular calcification, which acts as a central driver of CAVD. In particular, TNF and TGFβ1 emerged as top candidate genes regulated by epigenetic mechanisms. TNF expression was positively associated with CAVD risk and linked to enhancer demethylation (β_SMR = 0.212), consistent with its established role in promoting vascular inflammation and osteogenic transformation of valve interstitial cells [33–36]. Likewise, increased TGFβ1 expression (β_SMR **=** 1.0928) aligns with in vivo studies demonstrating its involvement in myofibroblast activation, extracellular matrix remodeling, and valve fibrosis [37–40]. These findings reinforce the dual contribution of pro-inflammatory cytokines and fibrotic signaling in CAVD progression. Another compelling candidate, TLR2, which plays a central role in innate immune activation [41–44], showed consistent associations in both blood and aortic tissue. This dual-compartment involvement points to systemic immune priming as a potential amplifier of localized valve inflammation.

Importantly, we also identified genes with protective or modulatory roles. PPARG, a nuclear receptor involved in lipid metabolism and anti-inflammatory signaling [45–47], was negatively associated with CAVD risk, suggesting a protective function. In contrast, elevated LTA expression and dysregulation of complement components C3 and C4A suggest broader disturbances in immune regulation. Notably, C4A demonstrated a protective effect, potentially through dampening complement activation, a mechanism increasingly recognized in cardiovascular inflammation [53–54]. Among the newly implicated pathways, AGER stood out due to its strong co-localization with inflammatory mediators such as IL-18, IL-17C, and CCL23 [48–52]. This supports its role in sustaining chronic inflammation within the valvular microenvironment, potentially linking innate immune sensing to downstream cytokine responses. To validate these findings in an independent population, we leveraged data from the UK Biobank. This large-scale cohort confirmed significant associations of the TNF (rs116454630) and TGFβ1 (rs34503210) loci with CAVD risk, and showed consistent directional trends for other key candidates, including TLR2, C4A, AGER, and IL6. These results provide robust population-level support for the genetic regulation of inflammatory processes in CAVD.

Taken together, our integrative analysis suggests that CAVD arises from a complex interplay between systemic immune dysregulation and local vascular inflammation. Despite its anatomically focal presentation, the disease appears to reflect widespread immune dysfunction [19,55–57]. The convergence of transcriptomic, genetic, and epigenetic evidence across multiple data types strengthens the credibility of our findings and underscores the potential for identifying biomarkers and therapeutic targets. Nonetheless, we acknowledge key limitations. Our study is based primarily on computational inferences, and experimental validation, particularly functional assays in human valve tissue, remains essential to confirm causality. Additionally, the relatively small number of aortic valve samples in available datasets limits the generalizability of some findings. Future work should focus on expanding tissue-specific datasets and exploring therapeutic modulation of implicated pathways.

## Conclusion

Our multi-omics analysis reveals that inflammation plays a central, genetically regulated role in calcific aortic valve disease (CAVD). We identified key inflammatory genes, including TNF, TGFβ1, AGER, C4A, TLR2, and PPARG, that are significantly associated with CAVD risk through integrated transcriptomic, genetic, and epigenetic analyses. Tissue-specific regulation of AGER, C4A, and TLR2 implicates local immune signaling in the aortic valve, while epigenetic upregulation of TNF and TGFβ1 supports their causal role in promoting inflammation and calcification. Protective associations for PPARG and C4A suggest counter-regulatory pathways that may modulate disease severity. Validation in the UK Biobank confirmed several of these associations, reinforcing their relevance at the population level. Collectively, our findings strengthen the concept of CAVD as an immune-mediated disease and highlight potential targets for biomarker development and therapeutic intervention.

## Acknowledgements

We thank all the investigators and subjects who participated in this project.

## Statements

### Ethics statement

The studies involving human participants were reviewed and approved by the Ethics Committees of the First Affiliated Hospital of Xinjiang Medical University. No informed consent was required.

### Author contributions

DSY, MYT, and AD conceptualized, designed, and supervised the study. DSY, RR collected the data and performed the data analysis. DSY, RR, LXM, and LXL wrote and revised the manuscript. KM, CJ, and LX organized and graphed the data. WX, YZX, and LF provided advice for the study and revised the manuscript for important intellectual content. LXL, AD revised the manuscript. The authors read and approved the final manuscript.

### Data availability statement

The datasets supporting the conclusions of this article will be made available by the authors without undue reservation.

## Abbrevations

CAVD: Calcific aortic valve disease
eQTL: expression quantitative trait loci
mQTL: DNA methylation QTL
SMR: summary data-based Mendelian randomization
DEGs: Differentially expressed genes
GWAS: genome-wide association study
VICs: valve interstitial cells
AS: atherosclerosis
CSEA: Cell-specific enrichment analysis

